# The Effect of Image Resolution on Automated Classification of Chest X-rays

**DOI:** 10.1101/2021.07.30.21261225

**Authors:** Md Inzamam Ul Haque, Abhishek K. Dubey, Jacob D. Hinkle

## Abstract

Deep learning models have received much attention lately for their ability to achieve expert-level performance on the accurate automated analysis of chest X-rays. Although publicly available chest X-ray datasets include high resolution images, most models are trained on reduced size images due to limitations on GPU memory and training time. As compute capability continues to advance, it will become feasible to train large convolutional neural networks on high-resolution images. To verify that this will lead to increased performance, we perform a systematic evaluation to measure the effect of input chest X-ray image resolution on accuracy. This study is based on the publicly available MIMIC-CXR-JPG dataset, comprising 377,110 high resolution chest X-ray images, and provided with 14 labels to the corresponding free-text radiology reports. Our original hypothesis that increased resolution would lead to higher accuracy held true for some but not all of the tasks. We find, interestingly, that tasks that require a large receptive field are better suited to downscaled input images, and we verify this qualitatively by inspecting effective receptive fields and class activation maps of trained models. Finally, we show that stacking an ensemble across resolutions outperforms each individual learner at all input resolutions while providing interpretable scale weights, suggesting that multi-scale features are crucially important to information extraction from high-resolution chest X-rays.

## 1 Introduction

Chest X-ray (CXR) is the most commonly used medical imaging modality in the world for the characterization and detection of cardiopulmonary conditions [1]. Timely analysis of CXRs is important to effective clinical care, yet delays and backlogs are a common scenario due to overburdened systems in many large healthcare centers or scarcity of radiologists in non-urban areas [2]. Moreover, to visually analyze such large-scale data requires expertise and concentration, and can be prone to bias [2]. Accurate automated analysis of CXRs would allow radiologists to improve the efficiency of their workflow on assessing abnormalities in CXRs X-rays and help extend this capability to under-served regions [3], [4].

In recent years, deep learning (DL) [5] has seen remarkable success in medical image analysis. In particular, deep convolutional neural networks (CNNs) have proven to be powerful tools to achieve and exceed state-of-the-art classification performance across image application domains [6]. CNNs excel in feature extraction with minimal data preprocessing, eliminating the need for manual curation requiring domain expertise.

Pioneering studies have been done in computer-aided diagnosis on chest radiographs focusing on a specific disease such as pulmonary tuberculosis classification [7], lung nodule detection [8], chronic obstructive pulmonary disease [9]. The recent release of the large-scale publicly available datasets have enabled many studies using deep learning for automated chest radiograph diagnosis [10], [11]. To increase the pre-trained DL models’ ability to generalize in clinical settings, some other studies have also been done, such as domain shift detection and removal to overcome heterogeneity in chest X-ray distributions [12], and fairness gaps in deep chest X-ray classifiers [13]. Tang et al. [14] compared classification performance of different deep CNNs with different input image sizes on the NIH ChestXray14 dataset. Recently, Bressem et al. [15] compared sixteen different architectures of CNN regarding chest radiograph classification performance on two openly available datasets, the CheXpert and COVID-19 Image Data Collection. However, there still exists a performance gap between these algorithms and radiologists for many categories, possibly due to the class imbalance of the dataset and label noise caused by natural language processing (NLP) [16], [17]. Nevertheless, studies for rapid review and reporting of abnormal chest X-rays can be galvanized with a pre-trained deep CNN.

The development of robust DL models is dependent on the availability of large quantities of training data. Fortunately, automated analysis of CXRs has seen considerable recent interest following the introduction of large publicly available chest radiograph datasets such as MIMIC-CXR [18], MIMIC-CXR-JPG [19], Chest-Xray8 [20], Chest-Xray14 [21], and CheXpert [22]. Leveraging these large-scale datasets, DL-based medical image classifiers have demonstrated nearly radiologist-level accuracy in diagnostic classification [23].

Most studies of DL-based automated chest X-ray abnormality classification have employed downscaling to reduce the native resolution to 256×256 pixels [23], or by dividing images into 256×256 pixel patches to extract local features from them [24]. But the aforementioned publicly available datasets come with high-resolution images, larger than 256×256 resolution. For example, the native resolution of chest X-ray images in the MIMIC-CXR-JPG dataset is 2500×3056 pixels mostly whereas the ChestXray14 dataset comes with 1024×0124 images.

In this study, we systematically evaluate the possibility of achieving higher clinical task performance by using high-resolution images instead of downscaled low-resolution images. We expect that increasing the image size will yield higher task performance for chest X-ray classification. To verify our expectation, we used a deep CNN model utilizing the MIMIC-CXR-JPG dataset, trained the CNN model with 256×256, 512×512, 1024×1024, and 2048×2048 resolution images, and then compared the task performances of different resolutions. As this is a multitask classification problem, we will see later in the results section that our expectation does not hold true for all tasks. We find that individual task performance is dependent on the effective receptive field of the task. Interestingly, tasks that require a large receptive field are better suited to downscaled input images, and vice versa. We verify this by inspecting effective receptive fields of the trained CNN models at each resolution and class activation maps of trained models for different tasks. Finally, we show that stacking ensemble across resolutions outperforms each individual learner at all input resolutions. We also provide interpretable scale weights of the ensemble model suggesting the need for multi-scale features for automated analysis of high-resolution chest X-rays.

## 2 Methods

We trained CNN models on downscaled CXRs and examined their performance as a function of downscaled resolution. Additionally, we examined the effective receptive field of these networks and their class activation mappings, and we compared them to a multi-scale stacked ensemble model. The following sections describe these components in more detail.

### 2.1 Downscaling

CNN models became popular after getting state-of-the-art performance in solving the ImageNet challenge in 2012. Since then, most CNN models performing image classification use pre-trained weights with ImageNet dataset [25]. ImageNet includes images with a resolution of 482×415 pixels or 469×387 pixels. After the introduction of AlexNet [6], cropping down images to 256×256 pixels has become a de-facto standard for CNN-based image classification.

Historically and even currently, people use low-resolution images for various reasons. Smaller images are easier to distribute and require less storage space than large, native-resolution images. Also, in many computer vision tasks, common objects can be discerned easily with a low-resolution image. Moreover, it is much easier and faster to train a deep CNN with low-resolution images. Although low-resolution images are common in the deep learning community, medical imaging data such as chest radiographs are often available in high resolution. Studies have shown that subtle radiological findings, such as hairline fractures, or small pneumothoraces, are less likely to be visible at lower resolutions. For example, Ribli et al. [26] showed lesions were most accurately detected in mammograms with 1700×2100 pixels images. Also, some top entries in the Society of Imaging Informatics in Medicine and American College of Radiology Pneumothorax Challenge such as [27] used input images with 1024×1024 pixel resolution. Datasets with high-resolution images are now publicly available such as OpenImages [28] which has roughly 6M images of size 1024×768 pixels and comes with labels. Therefore, such datasets can be used to train a deep CNN, and then the pre-trained weights can be used to detect subtle medical findings using CNN and it is an important research question. Although the common trend in medical image classification studies is to use low-resolution images, it may sacrifice important clinical information in the images, which are more clearly visible with high-resolution images. To test this hypothesis with the MIMIC-CXR-JPG dataset, we applied image downscaling from native resolution to 2048×2048 pixels, 1024×1024 pixels, 512×512 pixels, and 256×256 pixels, and then evaluated clinical task performance after training a CNN model using each of the downscaled image resolutions.

### 2.2 DenseNet Models

As most prior studies on CXR classification achieved the best result with a 121-layer DenseNet [29] model, it was adopted as the base model for this study. Initially, the model was trained from scratch, but we obtained a better performance later with pre-trained weights from ImageNet. The model is trained with a multi-label binary cross-entropy (BCE) loss. We normalize all the images via the mean and standard deviation of the ImageNet dataset. We use data augmentation techniques such as random horizontal flip and uniformly random rotation between -15 and 15 degrees.

Adam optimizer [30] is used with different initial learning rates for different image sizes. Learning rates are scaled with effective batch size for different image sizes as shown in Table 1. Learning rate decay is used such that it is multiplied by 0.5 if the validation loss does not decrease over three epochs. The training is stopped early if the validation loss doesn’t improve over ten epochs. Instead of using the official train-validation-test split that comes with the MIMIC-CXR-JPG dataset, five-fold cross-validation is used with 20% data as test data. The remaining 80% data is divided into a 90-10 split for training-validation data. Also, automatic mixed precision (AMP) is used which gave up to approximately 20% speedup in training which is crucial for training high-resolution images.

**Table 1:** Initial learning rate and effective batch size for different input image sizes. Batch sizes were chosen to maximize computational efficiency subject to GPU memory constraints then learning rates scaled proportional to batch size followed by manual adjustment.

| Input image size | Initial learning rate | Effective batch size |
| --- | --- | --- |
| 256x256 | 0.0005 | 512 |
| 512x512 | 0.00025 | 256 |
| 1024x1024 | 0.00016 | 168 |
| 2048x2048 | 0.00008204 | 42 |

### 2.3 Effective Receptive Field

The theoretical receptive field, or field of view, of a CNN is the collection of pixels of the input on which a pixel in the output features of the CNN depends on. Although, for fully connected networks, a unit depends on the whole input, for fully convolutional networks, each pixel’s receptive field can depend only on a region determined by the size and stride of convolutional and local pooling layers in the network. The receptive field is an important element for a deep CNN since areas in an input image outside the receptive field do not contribute to the model prediction. Therefore, it is important that the relevant areas in an input image are covered by the receptive field to get better predictions from the model.

We calculated the theoretical receptive field of the DenseNet121 model to be 2071×2071 pixels, which is larger than the four scales we are dealing with in this paper. It may seem seeing this big of a receptive field that all the different scales should have similar performance. In reality, this is misleading because the theoretical receptive field is not the most meaningful measure of receptive field as shown in [31]. It is shown that in practice only the central pixels of a theoretical receptive field contribute significantly to the output and hence for different scales this area of interest can be different which is discussed next.

The center pixels in a receptive field having the most impact on the output is called the effective receptive field (ERF). Luo et al. showed that the distribution of impact within the receptive field is asymptotically Gaussian, and the effective receptive field only takes a fraction of the theoretical receptive field [31]. Also, the effective receptive field was shown to get larger as the network is trained. The effective receptive field and its effect in deep CNNs is described in more detail in [31].

To compute the ERF, we load a trained model, set it in evaluation mode, load the data and compute the uncentered sample variance by averaging the square gradients. We take one gradient per image, using the center pixel and backpropagation. We also augment the data by large random translations when computing the ERF. We denote the value of image *x*_*i*_ at pixel (*p, q*) by *x*_*i*_(*p, q*) and channel *k*, pixel (*u, v*) of the neural network’s last convolutional layer activation by *f*^*k*^ (*x*_*i*_, *u, v*). The ERF is defined as a scalar image whose value indicates by how much each pixel *x*_*i*_(*p, q*) contributes to that activation. Impact is measured by the partial derivative 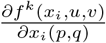. Note that this derivative depends not only on the weights of the neural network but also on the input image *x*_*i*_. The partial derivative 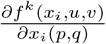 can be easily computed with back-propagation.

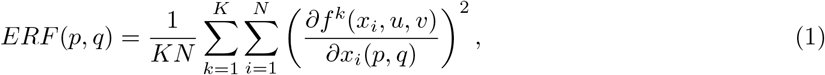

where *K* is the number of channels in the last convolutional layer of the network and (*u, v*) is the central point in each image *x*_*i*_. Because gradients can be both positive and negative in backpropagation we square the gradients first and average them to avoid cancellation, whereas in the original paper of ERF [31], the authors do not square the gradients when averaging.

We computed the ERF of trained models for different scale input images. Fig. 1 shows the comparison of ERF for DenseNet121 models trained with different resolution images. The ERF clearly decreases in size with respect to the image domain with increasing image resolution. This hints at the need for larger receptive fields when employing CNNs on high-resolution images.

**Figure 1:**
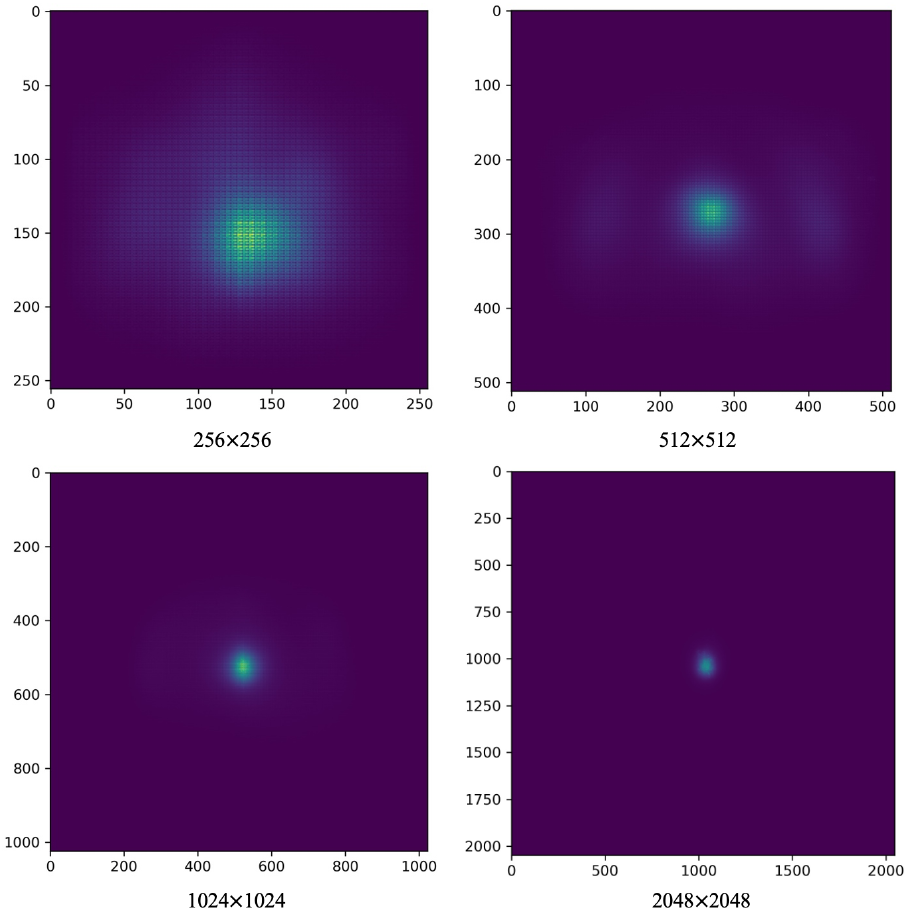
Effective receptive field of trained DenseNet121 models relative to image size, for different image resolutions. As the image resolution increases, ERF decreases due to corresponding decrease in pixel size.

### 2.4 Stacked scale-ensemble Model

To improve performance while retaining the ability to use pre-trained models, we employed a stacked ensemble model where the four models trained with different scale inputs (256×256, 512×512, 1024×1024, and 2048×2048) acted as learners. For a specific label, we use the concatenated label probabilities of the validation data from these four classifiers as features to train a particular logistic regression model to obtain weights for each scale. This ensemble technique is visualized in Fig. 2.

**Figure 2:**
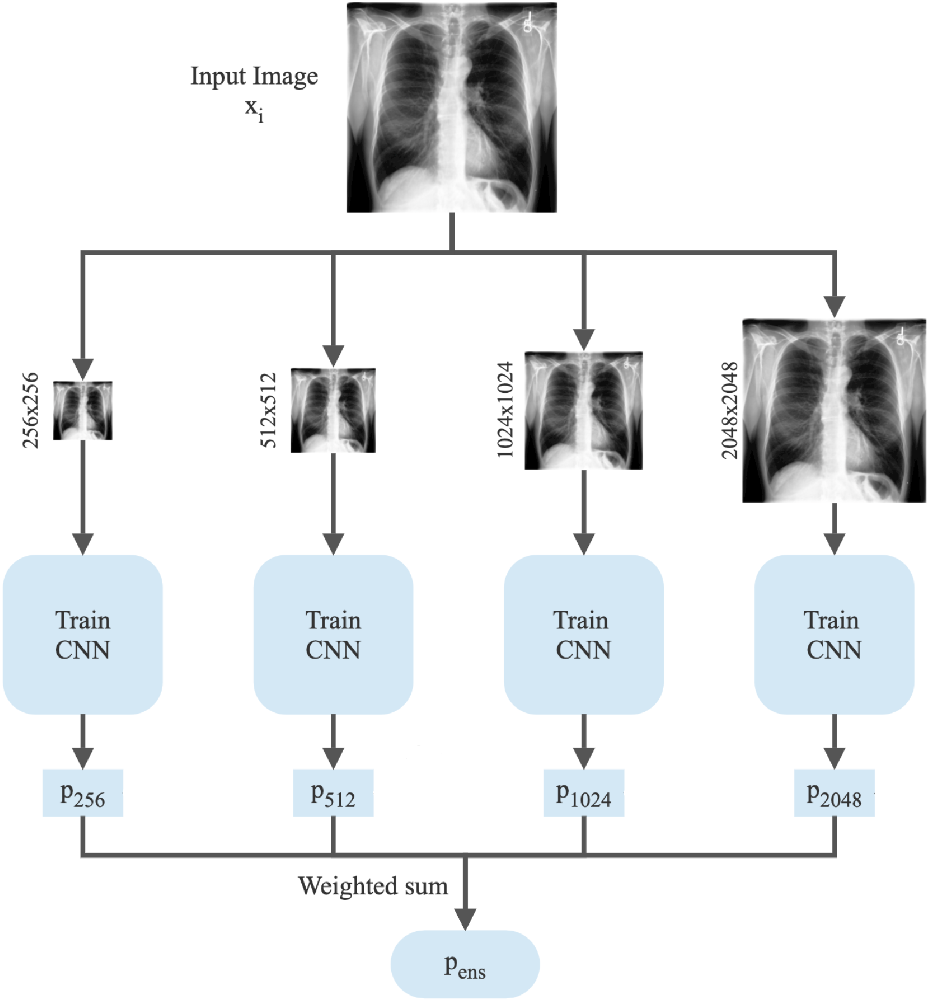
Workflow for the stacked ensemble model for a single input image. First, the image is resized to 256×256, 512×512, 1024×1024, and 2048×2048 resolution images and then passed to their corresponding fine-tuned models. The outputs from the models are then passed to a sigmoid function to get the output probabilities for each model. Finally, for each of the 14 labels, the predictions from the four models are multiplied by the learned weights to get the stacked ensemble prediction.

Our multi-label stacked ensemble model has a single parameter *w* ∈ ℝ^14×4^ which is a two-dimensional tensor such that the weight for finding *f* and scale *s* is *w*_*fs*_. We use a softmax parametrization to ensure that the learned weights of the four scales for a specific label sum to one:

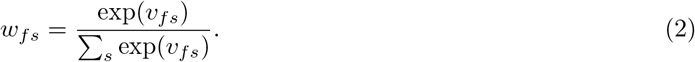

This parameterization satisfies ∑_*s*_ *w*_*fs*_ = 1 for each finding *f*. We initialize the weights *w*_*fs*_ as uniform by setting *v*_*fs*_ = 0. The weights are then used to average predicted label probabilities from each scale:

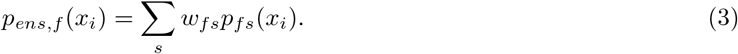

For an input image *x*_*i*_ with predicted logits *l*_*fs*_(*x*_*i*_), we implement a numerically stable computation of ∑_*s*_ *w*_*fs*_*σ*(*l*_*fs*_(*x*_*i*_)) for each finding *f*, where *σ*(*z*) = (1 + exp(-*z*))^*-*1^ is the sigmoid function. Note that each label *f* is treated separately in this model, even though all labels are computed at once in our implementation.

We train this stacking ensemble model with the input being the output logits of the validation data computed from the four classifiers of four scales. We use a learning rate of 0.1 that decays exponentially and we train the model for 100 epochs, verifying that the weights converge. After getting the learned weights from this model, we multiply them with the predictions computed from the test data to get the ensemble probabilities for each label.

We have also tried a simple voting ensemble model where we take the output probabilities from the test data for each scale and average them to get the ensemble prediction for each label. This voting ensemble model performed nearly as well as the stacked ensemble model but lacked the explainability that our stacked ensemble provides, which will be explained in the next section. The performance of the stacked ensemble model as well as the distribution of the learned weights of each label for different scales are discussed in the result section.

## 3 Results

We discuss the MIMIC-CXR-JPG dataset here in detail along with the computational environment used to train the CNN models, and execute the experiments. The performance for individual resolution model as well as the stacked ensemble model are presented. Finally, we visualize class activation maps using Grad-CAM.

### 3.1 Dataset

MIMIC-CXR-JPG dataset has been used in this study. It is a large dataset consisting of 377,110 chest X-rays associated with 227,827 imaging studies. This dataset was developed on a study level - a collection of images associated with a single report is referred to as a study. Therefore, a single study can have multiple chest X-rays with frontal and lateral views. There are a total of 2,43,334 frontal-view images and a total of 1,17,968 lateral-view images are present in the dataset.

We used all images in the dataset without adding any filter to exclude any image from our experiments. Also, we did not observe any kind of trend between frontal and lateral images in our experiments. Different image sizes are present in the dataset. Fig. 3 shows the distribution of image sizes in the MIMIC-CXR-JPG dataset. Here, we can see that the MIMIC-CXR-JPG dataset contains different size images, but as the histogram suggests, 2500×3056 pixels images by far the most common.

**Figure 3:**
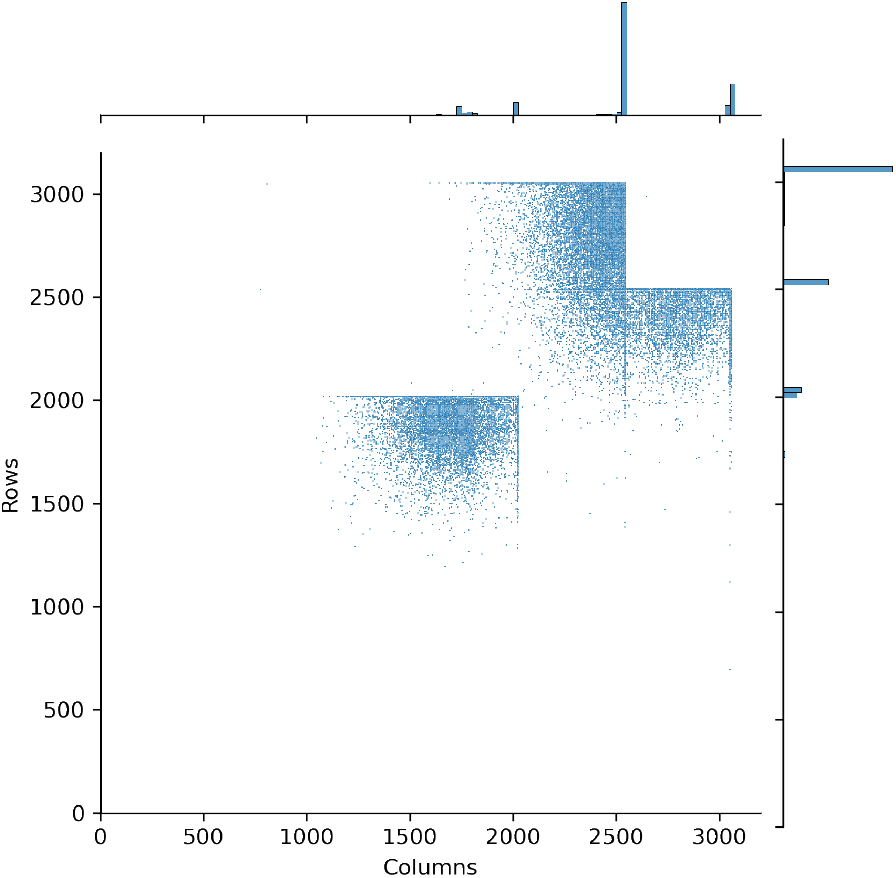
Distribution of 2D image sizes in the MIMIC-CXR-JPG dataset. Marginal distributions are shown as histograms along the top and right axes. The scatter plot shows many actual native resolutions, but the marginal histograms show that these are concentrated in a few common resolutions: approximately 2000×2000, 2500×3000, and 3000×2500.

The JPEG images in this dataset are converted from Digital Imaging and Communications in Medicine (DICOM) format. Although the DICOM format is commonly used in clinical practice and can store a large amount of meta-data, it can be difficult to comprehend for non-domain people. Apart from taking a lot less storage size, the JPEG images are also easy to comprehend. The images were de-identified and then exported to JPEG format. First, image pixels were extracted from the DICOM file, and then the pixel values were normalized to the range [0,255]. It was checked to see if pixel values were inverted and if necessary images were inverted to make sure that the highest pixel value appears white and the lowest pixel value appears black in the image. Contrast Limited Adaptive Histogram Equalization (CLAHE) was then applied to enhance the contrast of the images. Finally, the images were converted to the JPEG format with a quality factor of 95 and a bit-depth of 8-bit (0-255).

Chest X-ray images are provided with 14 labels - Atelectasis, Cardiomegaly, Consolidation, Edema, Enlarged Cardiomediastinum, Fracture, Lung Lesion, Lung Opacity, No Finding, Pleural Effusion, Pleural Other, Pneumonia, Pneumothorax, and Support Devices. These labels were derived from the impression section (83.2%), findings sections (12.2%), or final section (4.6%) of the radiology reports. Two open-source labeler tools, [32] and [22] were used to derive these 14 labels from a radiology report. Labels in this dataset have a high degree of missingness and come in four flavors: positive, negative, not mentioned (missing), and unknown. From different labeling methods outlined in [22], U-Zeros has been used in this paper where all non-positive labels including ‘negative’, ‘missing’, or ‘uncertain’ were aggregated into ‘negative’ label.

### 3.2 Computational Environment

NVIDIA DGX-A100 system has been used to train the DL models consisting of 8 NVIDIA A100 40 GB GPUs (Graphics Processing Units), each having high bandwidth memory (HBM2). This system has a total of 320 GB GPU memory with a maximum of 6.5 kW system power usage. With a system memory of 1 TB and internal storage of 15 TB, it uses Dual AMD Rome 7742 as its CPU (128 cores). A100 GPUs accelerate large-scale workloads when training in parallel by connecting multiple GPUs through NVLink. Note that, it is also possible to partition an A100 GPU into as many as seven GPU instances, having their own high-bandwidth memory. During training, 4/6 GPUs have been used depending on traffic on this shared machine. Pytorch framework and torchvision package has been used to write the code for our methods.

Memory used by a single GPU in gigabytes (GB) during training for each resolution is given in Table 2. As the resolution increases, memory usage increases as expected. It shows the computational capacity and power needed to run the experiments we executed. For 256×256 and 512×512 resolutions, we have used four A100 GPUs, but for 1024×1024 and 2048×2048 resolutions, we have used six A100 GPUs in parallel to make the training faster. As we double the image size, trainable parameters hence run-time increases and images per second decreases during validation.

**Table 2:**
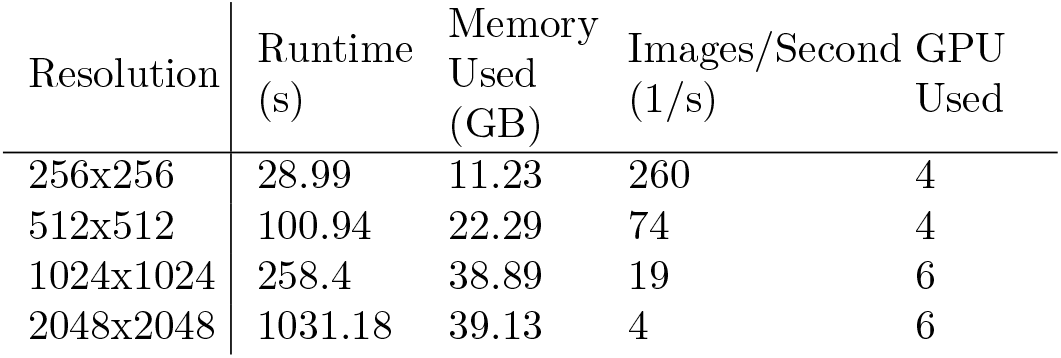
Runtime and images/sec for validation of DenseNet121 at each resolution along with the GPU memory used by each GPU during training on DGX-A100 system with eight 40GB GPUs.

### 3.3 Individual model performance

The area under the curve (AUC) score is used in this study to evaluate model performance. AUC measures the entire area underneath the receiver operating characteristic (ROC) curve. A model with higher AUC is better at predicting true positives and true negatives. AUC scores for all the 14 labels including their average are presented in Table 3 for different scales. As five-fold cross-validation was used during training, these AUC scores are the average of those five fold’s AUC scores and are shown along with standard deviations computed across the five folds. The highest value corresponding to any label is boldened in the Table 3. According to our hypothesis, the AUC score should increase for all the labels as we increase the input image size i.e., the 2048×2048 column of the table should have all the bold values. But as we can see from the table, this is not the case. Whereas some labels follow our expectations, some other labels don’t. For example, the AUC score of ‘Cardiomegaly’ decreases with increasing image size. A few labels tend to perform well with 512×512 resolution when increased from 256×256 resolution, but then their AUC scores decrease when increased further to 1024×1024 resolution.

**Table 3:**
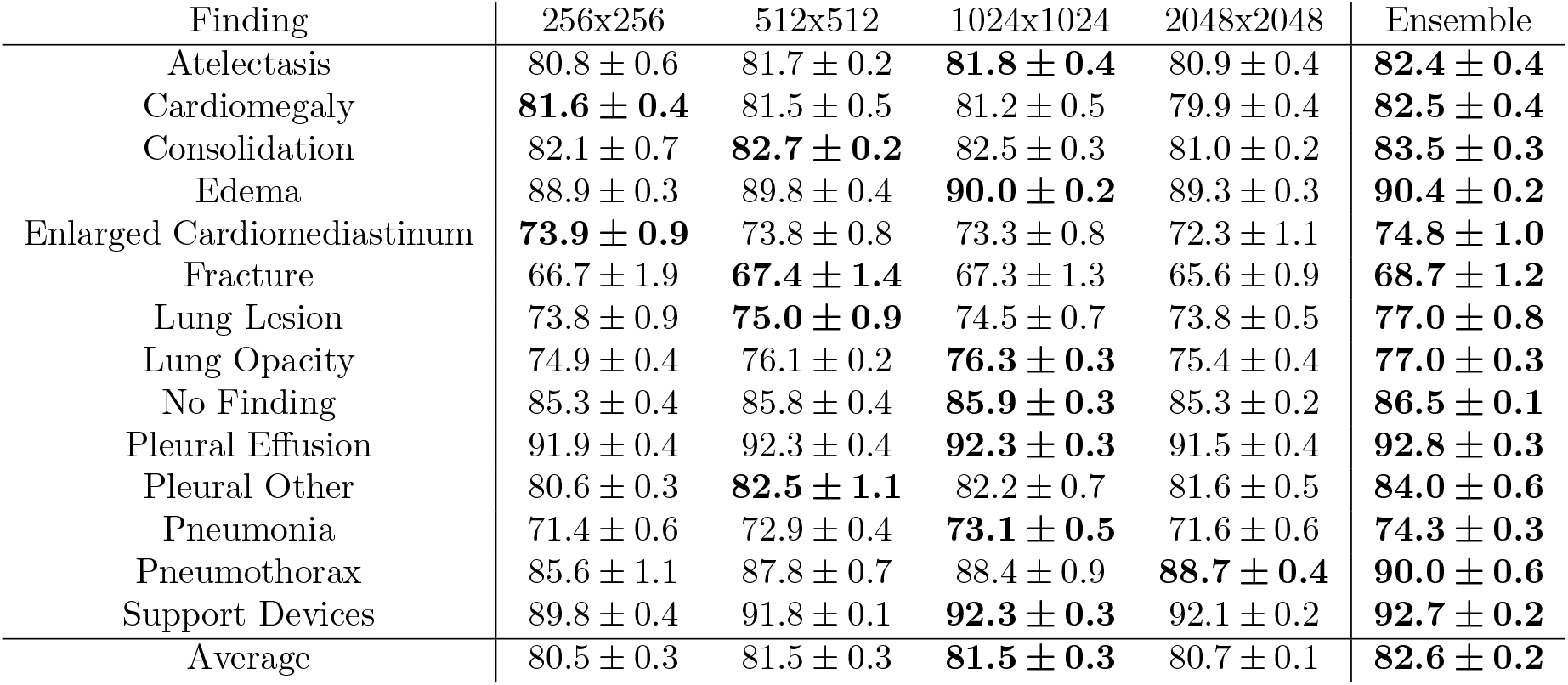
AUCs for densenet121 trained on downscaled images after ImageNet pretraining, in percent ± one standard deviation. Each scale was trained to predict all 14 labels in a 5-fold cross-validation split by patient. Each scale network outperforms all others for at least one task. Ensembling by simple probability averaging outperforms any individual scale network on every task simultaneously.

### 3.4 Visualization of Class Activation Maps

We suspect our result is due to the effective receptive field reduction with increasing input image resolution. As discussed before, it is important to have an ERF large enough so that no information is left out when making predictions. We have already seen that ERF decreases when image size increases. As a result, labels that take a larger portion in the input image such as ‘Cardiomegaly’, a condition of enlarged heart, tend to do well with reduced image size. This is because ERF is large enough for small-scale images to fit the enlarged heart inside the receptive field. As image size increases, ERF decreases, hence the receptive field can not capture the whole area of the heart and then make a bad prediction, which is exactly what we can see in Table 3. On the other hand, labels that are small in size, take a small area in the input image such as ‘Pneumothorax’, performs better as we increase the image size. This also makes sense because although the ERF is decreasing with increasing image size, it is still large enough to capture the small effective area of ‘Pneumothorax’. Large size objects can’t fit well in the small receptive field of high-resolution image networks and vice versa.

To test out and confirm the observation of the receptive field, we used Grad-CAM [33]. Grad-CAM is a technique for producing visual explanations for predictions made by a CNN model, which makes the model explainable and more transparent. Grad-CAM uses the gradients of any target label e.g. ‘Atelectasis’ or ‘Pneumonia’ in our study, flowing into the final convolutional layer to produce a coarse localization map highlighting the important regions in the image for predicting the label. We took the label ‘Cardiomegaly’, whose AUC score is the highest at the lowest image resolution (256×256), computed Grad-CAM for this label at different resolutions, as seen in Fig. 4. As seen in the figure, the network is not able to capture the entire heart when the resolution increases, hence the AUC score drops for this label as resolution increases. The bottom of Fig. 4 shows Grad-CAM visualizations for the label ‘Pneumothorax’ whose AUC score increases with increasing resolution. In this case, we find that the network is clearly focusing on the right portion of the image and predicting better as the resolution increases but has difficulty at low resolution, presumably due to inability to resolve fine features in the image.

**Figure 4:**
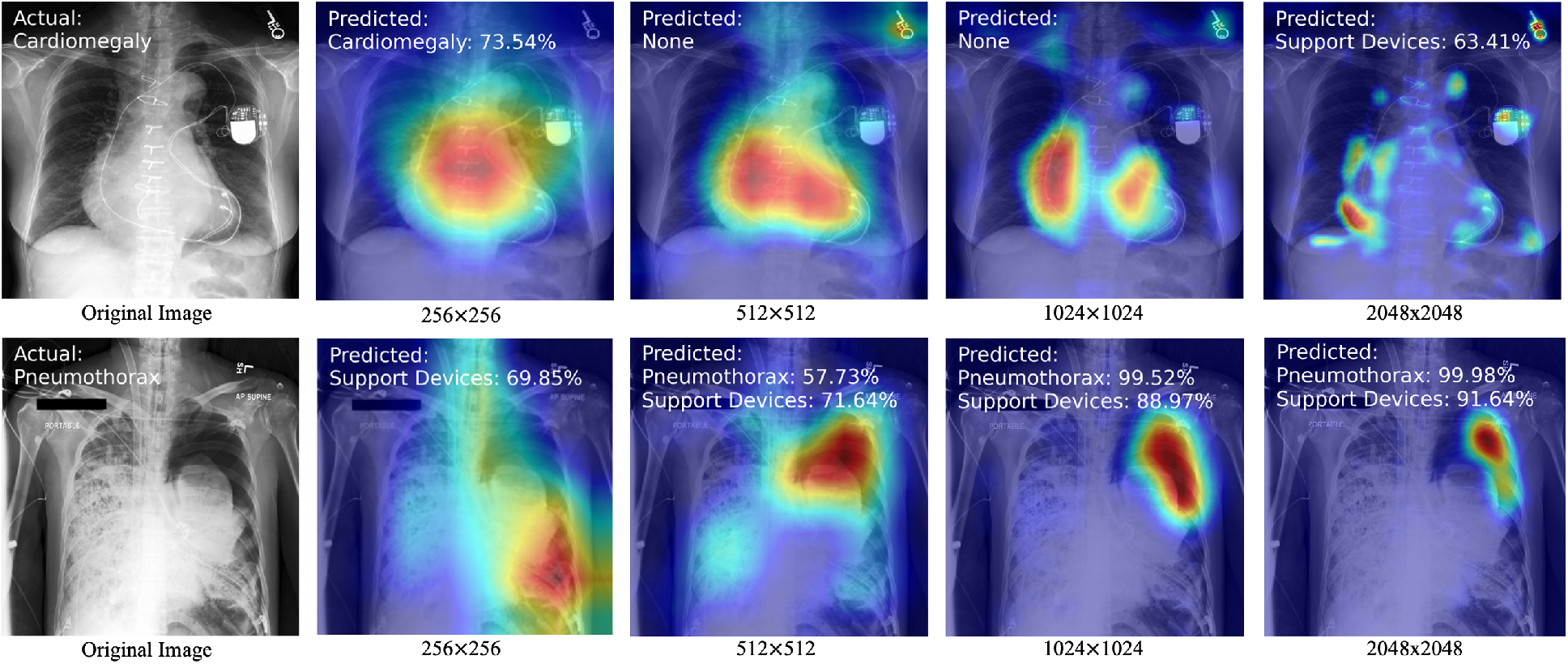
Grad-CAM computed to visualize influential regions for predicting ‘Cardiomegaly’ (top) and ‘Pneumothorax’ (bottom). Ground truth labels are shown overlaid on the original image, while for each Grad-CAM image, labels with probabilities greater than 50 percent are shown. The top row shows that ‘Cardiomegaly’ is only correctly predicted at 256×256 resolution, presumably due to effective receptive fields at high resolution being too small to encompass the entire heart. Conversely, the bottom row shows unreliable prediction of ‘Pneumothorax’ at coarse resolution due to lack of fine-scale information.

### 3.5 Ensemble Performance

As seen in Table 3, the ensemble model achieves the highest AUC for all 14 labels. For each label, the ensemble achieves an AUC at least 0.4% higher than the best-performing single-scale model, with an average AUC 1.1% higher than the 1024×1024 model. This gap indicates that even for any given label, specialization occurs across scales, with some examples better suited to different scales, which the ensemble is able to leverage to improve accuracy.

Fig. 5 shows the the distribution of the learned weight of the stacked ensemble model across four scales for each of 14 tasks. This bar chart shows which resolution is given more weights for a specific task when computing the prediction for the ensemble model. Note that the weights are shown in percentage and for a specific task, weights across the four scales sum to 100%. It is interesting to see that the learned weights resemble the AUC scores we got for individual models as shown in Table 3. For example, in computing prediction for ‘Atelectasis’ with the ensemble model, most weight (32.7%) has been given to the 1024×1024 scale, which is the highest performing individual model (bold) as seen from Table 3. In fact, this is true for ten out of the fourteen tasks.

**Figure 5:**
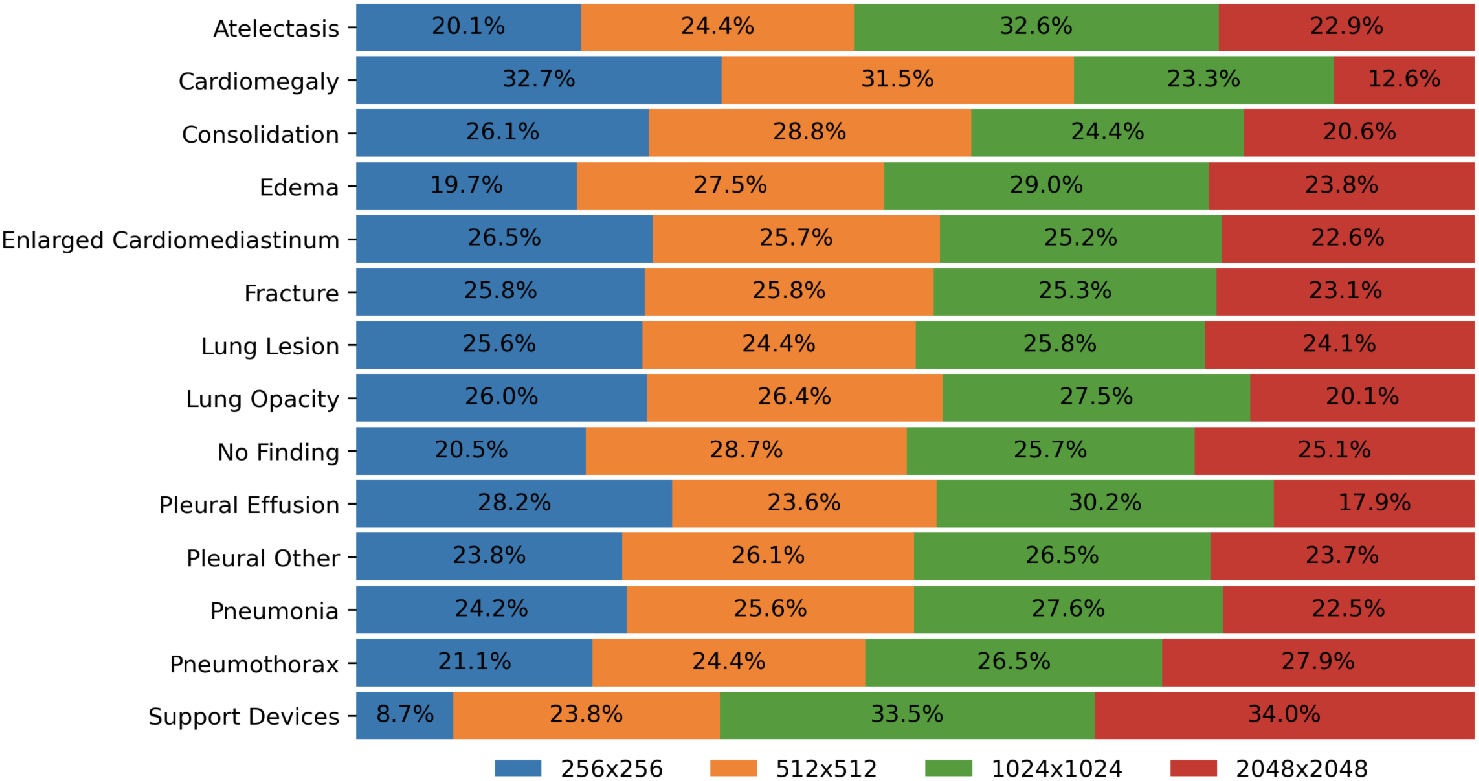
Barplot showing average stacked scale-ensemble weights for each resolution for each task. Tasks requiring mostly coarse-scale information, like Cardiomegaly, place larger weight on resolutions 256×256 and 512×512, while others such as Support Devices focus on higher resolutions.

## 4 Conclusion

In this work, we characterized the effect of downscaling on DenseNet performance for computer-aided detection using the MIMIC-CXR-JPG dataset. We found that of the 14 binary labeling tasks, some were better classified with low-resolution images whereas others were better classified with high-resolution images. We found the reason to be the ERF and its reduction with increasing image resolution. We also visualized class activation maps using Grad-CAM, showing that networks with improperly sized receptive fields do not focus on relevant areas in anatomy. This suggests using different networks for various tasks; for example ‘Cardiomegaly’ and ‘Enlarged Cardiomediastinum’ make better use of coarse-scale presentations while tasks requiring fine-scale information such as ‘Pneumothorax’ perform better with high-resolution images.

Additionally, we characterized an ensemble model using DenseNets trained with different scales and showed that it achieves the highest performance across all 14 labeling tasks. Further research can be performed to get even higher performance for this multitask classification problem.

We found that performance at all scales benefited from initializing our models with ImageNet pretrained weights. However, these ImageNet-pretrained weights were obtained by training on low-resolution (224×224 pixels) images. We suspect that pretraining on high-resolution image datasets containing millions of examples, like OpenImages or others, would further improve our results at high resolution.

This study shows the importance of image resolution and feature size in designing neural networks for automated radiological image classification. We have focused on DenseNet121 in this work due to the availability of pretrained weights, but new models with larger receptive fields could be used if pretraining at high resolution were feasible. For example, the feature pyramid network is known to have a large receptive field while incorporating coarse-scale information [34]. Given the success of our simple stacking approach, we expect future improvements in automated CXR classification to center on efficient extraction of high-resolution information without degrading the effective receptive field.

## Data Availability

This manuscript uses the MIMIC-CXR-JPG dataset, which is available to the public upon request.

https://physionet.org/content/mimic-cxr-jpg/2.0.0/

